# A Population-Based Cross-Sectional Investigation of COVID-19 Hospitalizations and Mortality Among Autistic People

**DOI:** 10.1101/2024.02.23.24303274

**Authors:** Dewy Nijhof, Filip Sosenko, Daniel Mackay, Michael Fleming, Bhautesh D Jani, Jill Pell, Chris Hatton, Deborah Cairns, Angela Henderson, Laura Ward, Ewelina Rydzewska, Maria Gardani, Elliot Millington, Craig Melville, the CVD-COVID-UK/COVID-IMPACT Consortium

**Author notes:** Joint first author (Dewy Nijhof, Filip Sosenko). Corresponding author: Dewy Nijhof.

## Abstract

**Background:** Current evidence suggests the possibility that autistic people may be at more risk of COVID-19 infection, hospitalisation, and mortality than the general population. Previous studies, however, are either limited in scale or do not investigate potential risk factors. Whilst many risk factors have been speculated to be responsible for severe COVID-19, this research has focused on general population samples.

**Methods:** Using data-linkage and a whole-country population, this study modelled associations between autism and COVID-19 hospitalisation and mortality risk in adults, investigating a multitude of clinical and demographic risk factors.

**Results:** Autistic adults had higher rates of hospitalisation, Standardised Incident Ratio 1.6 in 2020 and 1.3 in 2021, and mortality, Standardised Mortality Ratio 1.52 in 2020 and 1.34 in 2021, due to COVID-19 than the general population. In both populations, age, complex multimorbidity and vaccination status were the most significant predictors of COVID-19 hospitalisation and mortality. Effects of psychotropic medication varied by class.

**Conclusions:** Although similar factors exhibited a positive association with heightened risk of severe COVID-19 in both the autistic and general populations, with comparable effect sizes, mortality rates were elevated among the autistic population as compared to the general population. Specifically, the presence of complex multimorbidity and classification of prescribed medications may emerge as particularly significant predictors of severe COVID-19 among individuals within the autistic population due to higher prevalence of complex multimorbidity in the autistic population and variability in the association between medication classes and severe COVID-19 between both populations, though further research is needed.

## Introduction

In the UK, the earliest COVID-19 cases were identified in late January 2020 and a pandemic was officially declared in March 2020 (1). Increasing evidence suggests increased risk of susceptibility and severe disease progression of COVID-19 for certain conditions (2,3). For example, conditions such as diabetes and obesity were deemed risk factors (2). Subgroups within the population have been shown to be disproportionately at risk of COVID-19 but the full extent of these risk factors is unclear (4). Most research has focused on ethnic subpopulations or high-risk patient groups, but little is known about the risks of COVID-19 for autistic people^1^.

Autism Spectrum Disorder (ASD) is defined as a neurodevelopmental disorder. Its main characteristics are differences in social communication and interaction and repetitive and restrictive behaviours, including differences in sensory preferences (5). Whilst several genetical and immunological characteristics of autism have been hypothesised to affect COVID-19 infection, severity, and mortality risk (6–8), few studies have focused on differences in the clinical profiles of autistic people and the general population. Both physical and mental health conditions, such as obesity (2,3), gastrointestinal symptoms (3) and mental health conditions (9,10) have been associated with increased COVID-19 infection, disease severity and mortality in the general population. Several of these conditions often co-occur alongside autism (11,12). Specifically, psychotic disorders and mood disorders are more commonly experienced by autistic people (12) and have been linked with increased risk of COVID-19 mortality in the general population (9). It is further argued by some that sensory difficulties experienced by autistic people can hinder their ability to adhere to COVID-19 measures, such as wearing personal protective equipment (PPE) or strict hygiene rules, thereby increasing the potential for exposure to sources of infection (13,14). Similarly, differences in communication and social isolation can impact the ability to understand and follow COVID-19 related advice and affect help-seeking behaviours and patient-clinician communication in relation to health (13,14).

Furthermore, long-term conditions and multimorbidity prevalence is often accompanied by polypharmacy, the use of multiple medicines, which has been shown to contribute to increased COVID-19 morbidity in the general population (15,16) but has not been investigated in autistic populations. Yet, polypharmacy and more particularly prescription of antipsychotics are prevalent both amongst autistic children (17) and autistic adults (18,19). Commonly prescribed medications, including antipsychotics, antidepressants, and anticholinergic medication, are known to have the potential to increase the risk of pneumonia through immunosuppressive and other protective mechanisms (20). Moreover, antipsychotics have been shown to exert anti-inflammatory effects and disrupt immune responses (21), which may contribute even further to adverse COVID-19 outcomes. Indeed, several studies (22,23) and a meta-analysis (9) found increased risk of COVID-19 mortality associated with exposure to antipsychotics in the general population. Furthermore, recent research has raised the issue of potential adverse outcomes due to drug-drug interactions of prescribed medications and COVID-19 pharmacotherapies (24,25).

Few studies have investigated the association between autism and COVID-19 and results are inconsistent. One study investigating 36 autistic children in a residential facility did not find higher prevalence nor severity of COVID-19 in autistic young adults, though it should be mentioned that study controls (n=35) were of older age than the autistic group (6). Further, this study did not differentiate between young adults with autism only and young adults with co-occurring autism and intellectual disabilities (ID). Previous research suggests that people with ID are at higher risk of severe COVID-19 (27,28) and thus results may be confounded. Moreover, facilities in which individuals live in close proximity have been found to increase infection rates (29), confounding results even further. A meta-analysis found increased risk of COVID-19 mortality in autistic people only in unadjusted estimates and no evidence of increased intensive care admissions, but did not separate autism from ID or other developmental disorders (9). In contrast, a population-based study found that autistic males over 16 years had higher COVID-19 hospitalisation rates after stratifying for age, gender and morbidities that have been shown to increase COVID-19 infection rates compared to the general population (30).

As such, there is conflicting evidence regarding the risk of COVID-19 infection and outcome severity amongst autistic people. To the best of our knowledge, no studies have investigated whether this risk extends to COVID-19 mortality. Moreover, whilst previous studies speculated on the underlying causes of increased risk, none have systematically studied clinical risk factors for COVID-19 in the autistic population. Using a country-wide sample, the current study aimed to investigate whether there are differences in COVID-19 hospitalisation and mortality rates between autistic people and the general population. Furthermore, this study aimed to explore the association between demographic and clinical risk factors and severe COVID-19 in the autistic population.

## Methods

### Study Design

This was a cross-sectional study using a whole-country population. Data was subdivided into two time periods: (1) including the month in which the first lockdown started in the UK, 1 March 2020, to 31 December 2020; (2) 1 January 2021 to 31 December 2021. This division was made due to the vaccination programme not coming into full effect until 2021. While the first vaccination was received on 8 December 2020, due to vaccination priorities and logistics it is reasonable to assume that the majority of the study population had not received a COVID-19 vaccination in 2020. However, by the end of 2021, a greater proportion of the population had likely been afforded the opportunity to receive at least one COVID-19 vaccination. Consequently, COVID-19 rates in 2021 may vary significantly from those in 2020.

### Data sources and definitions

This study was conducted on behalf of the CVD-COVID-UK/COVID-IMPACT Consortium (31) (coordinated by the BHF Data Science Centre). The following datasets were used: GPES Data for Pandemic Planning and Research (GDPPR), Hospital Episode Statistics (HES), COVID-19 Second Generation Surveillance System (SGSS), Vaccination Events (VE), Civil Registry Deaths from the ONS (ONS-D), NHS BSA Dispensed Medicine (NHS BSA). The GDPPR, HES, SGSS, VE and NHS BSA datasets are updated through the National Health Service (NHS) and processed by NHS England, after which the data is available for secondary uses, such as research. The ONS dataset is updated through the Office for National Statistics (ONS). Approvals were obtained from the research consortium and from the University of Glasgow Ethics Committee.

Long-term conditions (LTCs) were identified using a list of 35 common conditions in the general UK population (32) using phenotypes sourced from the Cambridge Multimorbidity Score, using the GDPPR and HES datasets (33). This list includes conditions that have previously been identified as risk factors for COVID-19 infection and severity, for example obesity, diabetes, and hypertension (2). The list includes both physical and mental conditions but excludes autism. Complex multimorbidity was defined by the presence of at least three of those 35 LTCs of which at least one was physical (34). Polypharmacy was defined as the concurrent use of five prescription medications or more (35). Polypharmacy, use of antipsychotics, antidepressants, anticonvulsants, and psychotropic medication were identified using British National Formulary (BNF) codes. Prescribing data was examined if it fell within the following parameters: dispensed within 15 days before the date of testing positive for COVID-19 (cut-off date) and within 240 days before this cut-off date (35).

In the context of this study, severe COVID-19 included those who had a positive COVID-19 laboratory test and were hospitalised for COVID-19 or died from COVID-19 within 28 days from the test or died without a positive test, but COVID-19 is listed as a cause of death on their death certificate (any diagnostic position).

### Population

The study covered the whole adult (i.e., over 18 years old) population of England who were alive on 1 January 2020 and had at least one primary care record (for the purpose of establishing their autism status). Autistic individuals were identified using SNOMED CT diagnosis codes in the primary care dataset (GDPPR). Some evidence suggests that people with ID have a larger risk of severe COVID-19 (27,28). Further, research suggests that there may be complex interactions at play in relation to mortality and causes of mortality when it comes to comorbid autism-ID (36). As such, this study opted to investigate severe COVID-19 in autism only. Participants who had diagnoses for both autism and ID were included in the comparison group to control for potential confounding (having ID is a major risk factor for COVID-19 hospitalisation or death (27). Although there is a possibility that this may marginally reduce any differences found between autistic people and the comparison group, we maintain the stance that as a result of this decision, any inferences drawn from this study will be more generalisable to autistic people without intellectual disabilities-which remains the main focus of the study.

The whole population consisted of 180,860 (of which 28.5% female) autistic people and 48,383,120 (of which 50.4% female) people in the comparison group, hereafter referred to as general population. Age distribution in the two populations differed significantly, with a median age of 47 [interquartile range (IQR) 30] years in the general population and 26 [IQR 12] in the autistic population (t-test, *p*-value <0.001). This corresponds to known low levels of identification or recording of autism in primary care records for people aged over 30 years old (37) as well as diagnostic advances and autism awareness over the years (38). Demographic and health characteristics of the study participants are shown in *Table 1.* Characteristics for the 35 long-term conditions for both the whole population and analysis sample-(i.e., those infected with and hospitalised for COVID-19) can be found in the *Appendix*.

**Table 1.** Demographic and health characteristics of autistic and general population people hospitalised with a registered positive COVID-19 test.

|  | General population<br>(n = 8,425,470) | Autistic population<br>(n = 32,375) |  |  |  |
| --- | --- | --- | --- | --- | --- |
|  |  | Adjusted | P-value | Unadjusted | P-value |
| Age [Mdn., IQR] | 41.8 [30.0, 55.0] | - | - | 24.9 [21.0, 31.5] | <0.001 |
| Female (%) | 54.0 | 33.7 | <0.001 | 33.4 | <0.001 |
| Mean IMD decile (SD) | 5.4 (2.9) | 5.0 (2.8) | <0.001 | 5.1 (2.9) | <0.001 |
| *Ethnicity (%)* |  |  | <0.001 |  | <0.001 |
| White | 82.7 | 93.5 |  | 92.0 |  |
| Asian | 9.2 | 2.2 |  | 2.4 |  |
| Black | 3.7 | 1.6 |  | 2.0 |  |
| Mixed | 1.9 | 1.6 |  | 2.4 |  |
| Other | 2.4 | 1.1 |  | 1.3 |  |
| *Vaccine* |  |  | 0.8 |  | <0.001 |
| AstraZeneca | 21.2 | 23.4 |  | 30.1 |  |
| Moderna | 3.4 | 3.1 |  | 7.6 |  |
| Pfizer | 26.4 | 24.4 |  | 62.2 |  |
| *Vaccine doses* |  |  | >0.9 |  | <0.001 |
| 0 | 49.3 | 49.1 |  | 50.8 |  |
| 1 | 18.3 | 18.4 |  | 19.4 |  |
| 2 | 32.5 | 32.5 |  | 29.8 |  |
| *Total number of prescribed medications* |  |  | <0.001 |  | <0.001 |
| 0 | 55.8 | 36.1 |  | 53.4 |  |
| 1-3 | 23.0 | 24.4 |  | 25.8 |  |
| 4-6 | 9.3 | 14.5 |  | 10.4 |  |
| 7-9 | 4.9 | 9.0 |  | 4.9 |  |
| 10+ | 7.0 | 16.1 |  | 5.4 |  |
| Mean prescription medications (SD) | 2.9 (4.5) | 5.1 (6.0) | <0.001 | 2.7 (3.6) | 0.002 |
| Polypharmacy (%) | 21.2 | 39.5 | <0.001 | 20.8 | 0.14 |
| *Medications split by class* |  |  | <0.001 |  | - |
| On Psychotropics (%) |  |  |  |  |  |
| No | 80.9 | 49.7 |  | - |  |
| Yes | 19.1 | 50.3 |  | - |  |
| Antipsychotics (%) | 1.3 | 12.7 | <0.001 | - | - |
| Antidepressants (%) | 17.1 | 45.1 | <0.001 | - | - |
| Anticonvulsants (%) | 3.9 | 11.7 | <0.001 | - | - |
| **Morbidity** |  |  |  |  |  |
| Mean LTCs (SD) | 1.0 (3.0) | 2.4 (4.0) | <0.001 | 1.2 (2.7) | <0.001 |
| Multimorbidity (%) | 11.9 | 24.3 | <0.001 | 12.4 | <0.001 |

From this whole population, participants were included in the analysis if they met at least one of two criteria: (1) they had a registered positive COVID-19 laboratory test (identified through GDPPR) or (2) COVID-19 was listed as a cause of death (either primary or other contributing causes using ICD-10 code U07.1 or U07.2) on their death certificate. In the case of multiple positive tests, the first instance was used. Information about LTCs, vaccination status, and prescribed medications were extracted using the time of positive test result.

### Data Analysis

Data was analysed within NHS England’s Secure Data Environment service for England, made available via the British Heart Foundation Data Science Centre’s CVD-COVID-UK/COVID-IMPACT Consortium (31), using a combination of Python (v3.7), SQL, and R (v4.03) scripts. The analysis was performed according to a pre-specified analysis plan published on GitHub (39), along with the phenotyping and analysis code.

Demographic (age, sex, ethnicity, Index of Multiple Deprivation (IMD)) and health characteristics (LTCs, complex multimorbidity, vaccination status, prescribed medications) were explored and compared using means, standard deviations, and t-tests for continuous data and frequency, percentages, and χ^2^-tests for categorical data. Standardised Incident Ratios (SIRs) and Standardised Mortality Ratios (SMRs) were calculated for 2020 and 2021 separately. SIRs were also calculated for hospitalisation (while positive for COVID-19) and severe COVID-19 (hospitalisation or death due to COVID-19). A binary logistic regression model was used to model prediction of severe COVID-19 infection using demographic and health factors as predictors. Note that modelling of antipsychotic, antidepressant, and anticonvulsant medication did not control for diagnosis of related mental health conditions due to the possibility of prescription of these medications for unrelated issues. There is evidence to suggest that overprescription, particularly of psychotropic medications, is a significant issue in relation to autistic adults (37,40) with and without psychiatric conditions (41). Further, though research on these medication classes in autistic people is scarce, limited evidence suggests variable effects regarding efficacy and tolerance of psychotropic medication in autistic people (41).

Further hierarchical logistic modelling was used to assess the contribution of each predictor category. Blinder-Oaxaca decomposition analysis was applied to decompose the mean difference in the risk of severe COVID-19 between the two population groups for each of the predictor categories used in the regression modelling (42).

Logistic regression was chosen as the main modelling technique rather than survival analysis due to its allowance for a larger number of records to be included. (Individuals who had a PCR test already in hospital, and those who died without a PCR test, cannot be included in survival analysis). Still, survival analysis in the form of a Cox proportional hazards model was conducted as a sensitivity test of the logistic modelling.

## Results

### Demographics

A total of 32,375 autistic people and 8,425,470 general population individuals were hospitalised with a positive COVID-19 test. Table 1 shows the demographic and health characteristics of autistic people and the general population used in the analysis (i.e., tested positive for COVID-19). Due to the younger age structure in the autistic population, both unadjusted and age-adjusted values are shown. The autistic group had a significantly different population structure, demonstrated by a lower percentage of females, higher chance of living in a deprived area (evidenced by lower mean IMD decile) and a higher percentage of white ethnicity. Autistic adults were equally likely as the general population to be vaccinated for COVID-19. Polypharmacy was more prevalent in the autistic population, with the mean numbers of prescription medications being 5.1 (SD 6.0) compared to 2.9 (SD 4.5) for the general population. Similarly, use of psychotropic, antipsychotic, antidepressant and anticonvulsant medications, was higher within the autistic population.

Autistic adults were also more likely to experience complex multimorbidity. From the list of 35 most common conditions in the UK, only three were not significantly higher for autistic adults (bronchiectasis, chronic kidney disease, and multiple sclerosis; see Appendix for full list). A few conditions stood out due to particularly higher prevalence in the autistic population: stroke/transient ischaemic attack, schizophrenia/bipolar disorder, psychoactive substance misuse, Parkinson’s disease, diabetes, depression, dementia, asthma, anxiety/other neurotic disorders, and anorexia/bulimia.

### Incidence, mortality and survival analysis

Table 2 shows the SIRs for autistic adults who were hospitalised due to COVID-19 or who classify as having severe COVID-19 (i.e., taking together COVID-19 hospitalisations and COVID-19 deaths). The ratios show a higher incidence of both hospitalisation and severe COVID-19 for autistic adults in comparison to the general population, although a decline over time is visible from 2020 to 2021.

**Table 2.** Standardised Incidence Ratios for autistic adults with general population as reference.

|  | COVID-19 Hospitalisation |  |  |  | Severe COVID-19 |  |  |  |
| --- | --- | --- | --- | --- | --- | --- | --- | --- |
|  | Observed | Expected | Excess | SIR [95% CI] | Observed | Expected | Excess | SIR [95% CI] |
| Mar-Dec 2020 | 372 | 234 | 138 | 1.6 [1.4, 1.7] | 382 | 244 | 138 | 1.6 [1.4, 1.7] |
| Jan-Dec 2021 | 603 | 461 | 142 | 1.3 [1.2, 1.4] | 620 | 469 | 151 | 1.3 [1.2, 1.4] |

Table 3 shows the Age Standardised Mortality Rate (ASMR) and Standardised Mortality Rates (SMR). ASMR takes the European Standard Population 2013 as reference and SMR takes the general population from the study sample as reference. Whilst ASMRs were higher for both groups within the study sample, the autistic population had particularly high mortality rates compared to the European Standard Population. Within the study sample, mortality rates for autistic adults were similarly increased compared to the general population. Similar to incidence rates, a decline from 2020 to 2021 is visible.

**Table 3.** Age Standardised Mortality Rates (ASMR) for the adult autistic and general population in study sample as compared to the European Standard Population 2013. Second column shows Standardised Mortality Ratios (SMR) for the autistic population as compared to the general population for the study sample only. Observed, expected and excess mortality for SMRs are shown.

|  | ASMR [95% CI] |  | SMR [95% CI] |  |  |  |
| --- | --- | --- | --- | --- | --- | --- |
|  | Autistic population | General population | Autistic population | Observed | Expected | Excess |
| Mar-Dec 2020 | 278.9 [278, 278] | 166.2 [166, 166] | 1.52 [1.04, 2.0] | 39 | 25.6 | 13.4 |
| Jan-Dec 2021 | 271.5 [271, 271] | 143.1 [143, 143] | 1.34 [0.93, 1.74] | 42 | 31.5 | 10.5 |

### Modelling of risk factors

Logistic regression modelling has been employed to investigate risk factors. The final model included two versions sharing core predictor variables and differed only in the aspect of medications. Model A focused on polypharmacy while model B focused on psychotropic medication.

Table 4 shows the results from the logistic regression models A and B. For both the autistic and general population, several factors were positively associated with increased risk of severe COVID-19 with similar effect sizes: age, ethnicity, complex multimorbidity (count of long-term conditions), polypharmacy (count of prescription medication), and antipsychotic prescription. Interestingly, the effect of complex multimorbidity varies by the extent of polypharmacy and vice versa. The effect of either risk factor is stronger when the score for the other is low. Again, effect sizes were similar between the two groups.

**Table 4.**
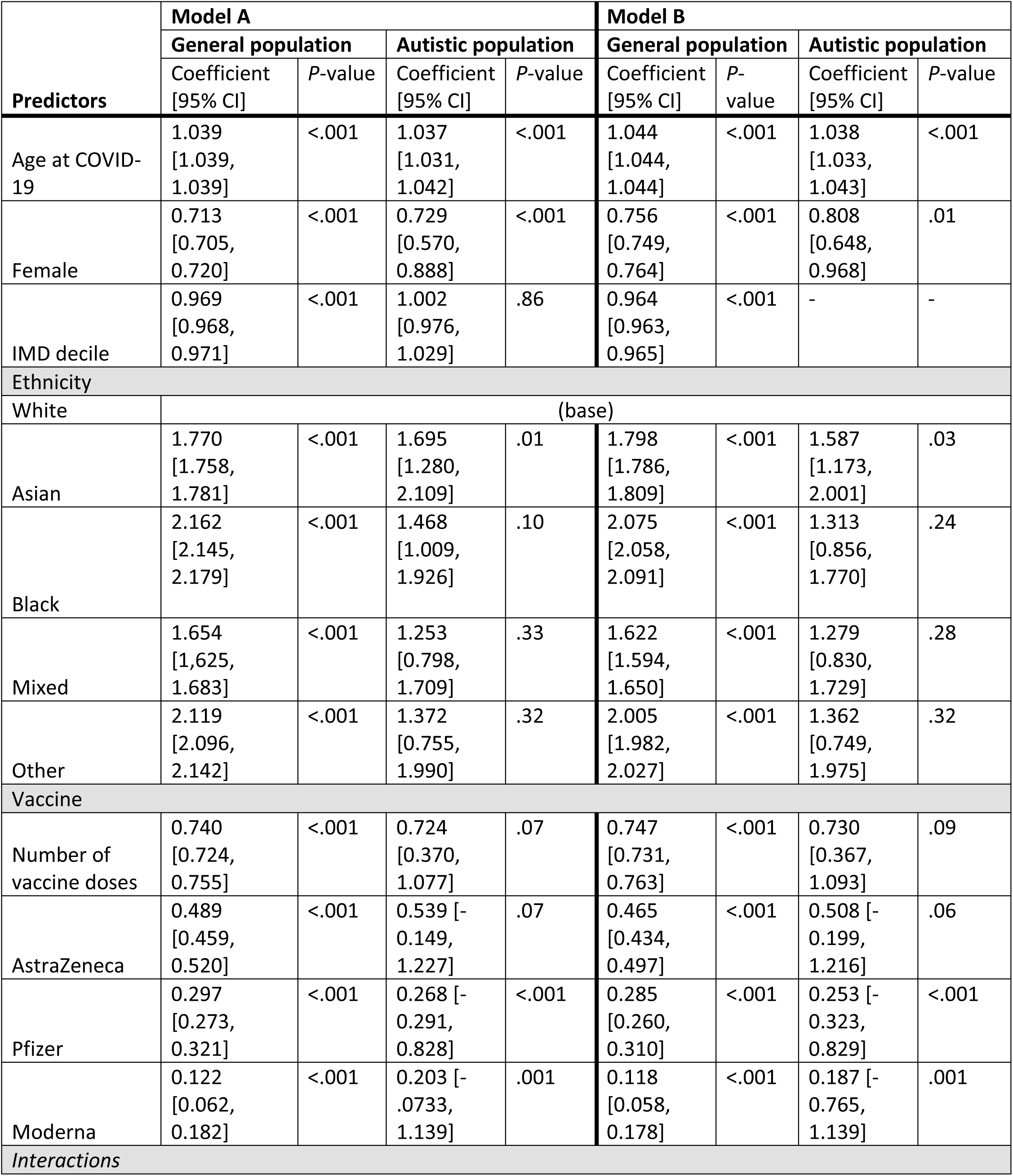
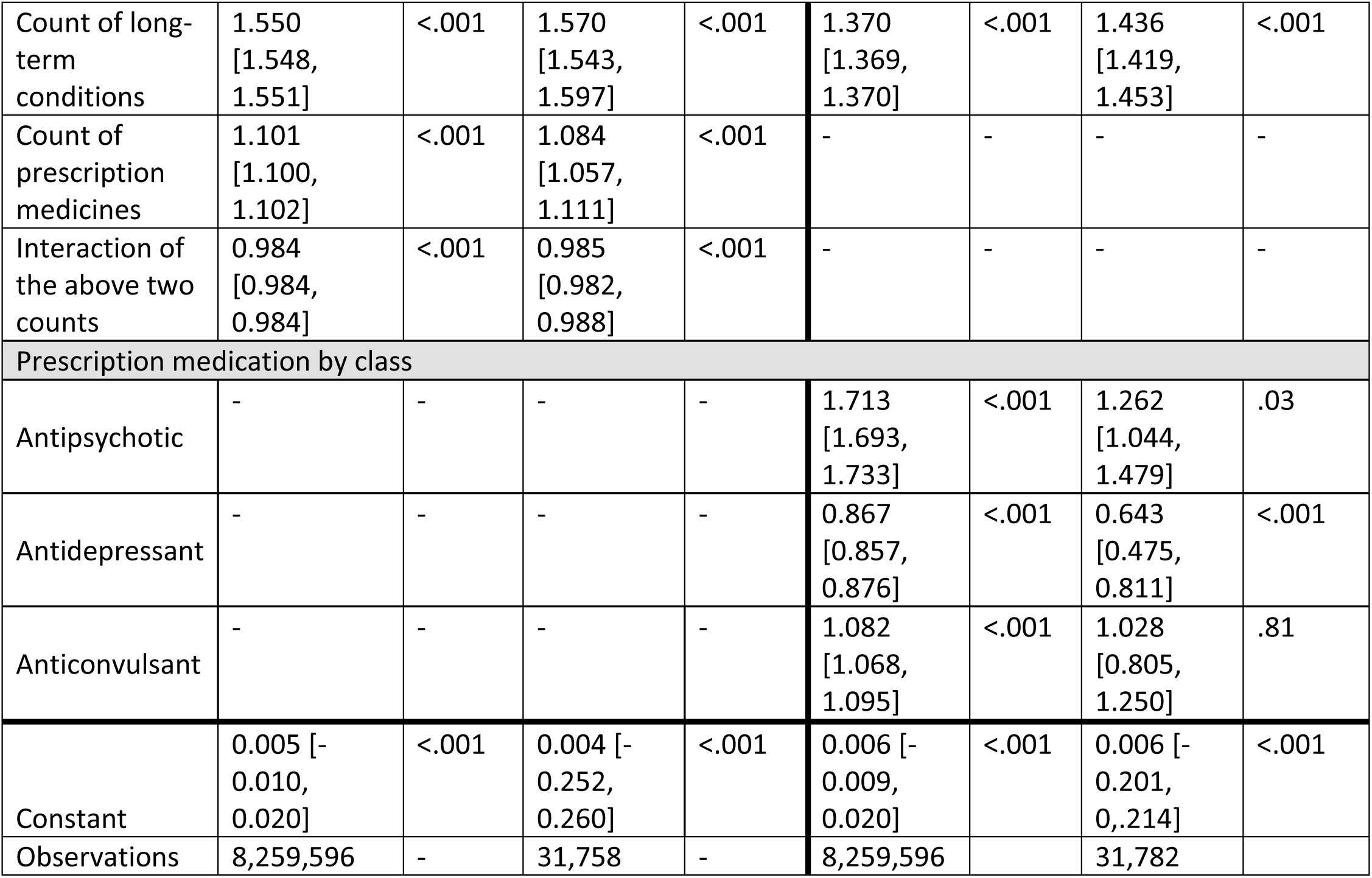
Results from the logistic models A and B fit to the autistic and general population separately.

Anticonvulsant prescription was significantly but weakly positively associated with increased risk for the general population only. Whereas Asian/Asian British ethnicity was the only ethnicity at risk for the autistic population, Black/Black British ethnicity was most at risk for the general population followed by Other, Asian/Asian British and Mixed ethnicity. Additionally, the effects of ethnicity and antipsychotic prescription on severe COVID-19 risk were more profound for the general population. Effects of long-term condition count were more pronounced for the autistic population. Contrastingly, female sex, Pfizer and Moderna vaccination, and antidepressant prescription were associated with decreased risk of severe COVID-19 for both the autistic and general population. Number of COVID-19 vaccination doses and AstraZeneca vaccination were significantly associated with decreased risk for the general population only. Antidepressant prescription effects were more pronounced for the autistic population. Table 5 shows the results from the survival analysis showing similar trends in associations, thus supporting findings from the logistic regression models.

**Table 5.** Results from the Cox proportional hazards model for model A and B.

| **Model A** |  |  |  |  |  |  |  |  |
| --- | --- | --- | --- | --- | --- | --- | --- | --- |
| **Variables** | **General population** |  |  |  | **Autistic population** |  |  |  |
|  | **β** | **SE** | **Exp(β) [95% CI lower, upper]** | **P-value** | **β** | **SE** | **Exp(β) [95% CI lower, upper]** | **P-value** |
| Age at COVID-19 | .03 | .0001 | 1.03 [1.03, 1.03] | <.001 | .03 | .003 | 1.03 [1.02, 1.03] | <.001 |
| Female | -.37 | .003 | .69 [.69, .69] | <.001 | -.28 | .087 | .75 [.63, .89] | .001 |
| IMD decile | -.03 | .0006 | .97 [.97, .98] | <.001 | - |  |  |  |
| **Ethnicity** |  |  |  |  |  |  |  |  |
| White |  |  |  |  | (base) |  |  |  |
| Asian | .51 | .005 | 1.67 [1.65, 1.69] | <.001 | .76 | .215 | 2.13 [1.40, 3.24] | <.001 |
| Black | .59 | .008 | 1.81 [1.78, 1.84] | <.001 | .21 | .240 | 1.24 [.77, 1.98] | .37 |
| Mixed | .39 | .014 | 1.47 [1.43, 1.51] | <.001 | -.14 | .282 | .87 [.50, 1.51] | .61 |
| Other | .59 | .011 | 1.81 [1.77, 1.85] | <.001 | .01 | .337 | 1.01 [.52, 1.95] | .98 |
| **Vaccinations** |  |  |  |  |  |  |  |  |
| Number of doses | -.23 | .008 | .80 [.78, .81] | <.001 | -.48 | .205 | .62 [.41, .92] | .01 |
| AstraZeneca | -.48 | .015 | .62 [.60, .64] | <.001 | -.09 | .394 | .92 [.42, 1.99] | .83 |
| Pfizer | -.95 | .012 | .39 [.38, .40] | <.001 | -1.03 | .314 | .36 [.19, .66] | .001 |
| Moderna | -1.93 | .034 | .15 [.14, .16] | <.001 | -2.04 | .748 | .13 [.03, .56] | .006 |
| **Interactions** |  |  |  |  |  |  |  |  |
| Count of long-term conditions | 0.22 | .0006 | 1.25 [1.25, 1.25] | <.001 | .35 | .012 | 1.42 [1.38, 1.45] | <.001 |
| Count of prescription medicines | .03 | .0006 | 1.03 [1.03, 1.03] | <.001 | .09 | .015 | 1.09 [1.06, 1.12] | <.001 |
| Interaction of the above two counts | -.006 | .00006 | .99 [.99, .99] | <.001 | -.01 | .002 | .99 [.99, .99] | <.001 |

| Model B |  |  |  |  |  |  |  |  |
| --- | --- | --- | --- | --- | --- | --- | --- | --- |
| Variables | General population |  |  |  | Autistic population |  |  |  |
|  | β | SE | HR [95% CI lower, upper] | P-value | β | SE | HR [95% CI lower, upper] | P-value |
| Age at COVID-19 | .025 | .0001 | 1.03 [1.03, 1.03] | <.001 | .03 | .002 | 1.03 [1.02, 1.03] | <.001 |
| Female | -.35 | .003 | .70 [.70, .71] | <.001 | -.19 | .086 | .83 [.70, .98] | .03 |
| IMD decile | -.03 | .001 | .97 [.97, .97] | <.001 | - |  |  |  |
| Ethnicity |  |  |  |  |  |  |  |  |
| White | (base) |  |  |  |  |  |  |  |
| Asian | .49 | .005 | 1.64 [1.62, 1.65] | <.001 | .77 | .215 | 2.14 [1.41, 3.28] | <.001 |
| Black | .57 | .008 | 1.77 [1.74, 1.79] | <.001 | .17 | .241 | 1.19 [.74, 1.90] | .48 |
| Mixed | .37 | .014 | 1.45 [1.41, 1.49] | <.001 | -.05 | .282 | .95 [.54, 1.65] | .85 |
| Other | .58 | .011 | 1,78 [1.75, 1.82] | <.001 | .11 | .337 | 1.12 [.58, 2.16] | .75 |
| Vaccinations |  |  |  |  |  |  |  |  |
| Number of doses | -.22 | .008 | .80 [.79, .81] | <.001 | -.45 | .203 | .64 [.43, .95] | .03 |
| AstraZeneca | -.48 | .015 | .62 [.60, .64] | <.001 | -.15 | .390 | .86 [.40, 1.85] | .70 |
| Pfizer | -.95 | .012 | .39 [.38, .40] | <.001 | -1.12 | .314 | .33 [.18, .60] | <.001 |
| Moderna | -1.95 | .034 | .14 [.13, .15] | <.001 | -2.13 | .750 | .12 [.03, .52] | .004 |
| LTCs |  |  |  |  |  |  |  |  |
| Count of long-term conditions | .17 | .0004 | 1.19 [1.19, 1.19] | <.001 | .28 | .008 | 1.32 [1.30, 1.34] | <.001 |
| Prescribed medications by class |  |  |  |  |  |  |  |  |
| Antipsychotic medication | -.01 | .007 | .99 [.97, 1.0] | .07 | .02 | .115 | 1.02 [.82, 1.28] | .85 |
| Antidepressive medication | -.17 | .004 | .85 [.84, .85] | <.001 | -.24 | .086 | .78 [.66, .93] | .005 |
| Anticonvulsive medication | .04 | .005 | 1.04 [1.03, 1.05] | <.001 | .11 | .111 | 1.12 [.90, 1.39] | .30 |

Hierarchical logistic modelling (Table 6) indicated that age, vaccination status and LTC count were the most important predictors for modelling severe COVID-19 infection in both autistic adults and the general population.

**Table 6.** Results from the hierarchical logistic regression showing Pseudo R^2^, Akaike Information Criterion (AIC) and Bayesian Information Criterion (BIC). Vaccine status is defined by the presence or absence of at least one COVID-19 vaccination.

|  | **Autistic population** |  |  | **General population** |  |  |
| --- | --- | --- | --- | --- | --- | --- |
|  | Pseudo R2 | AIC | BIC | Pseudo R2 | AIC | BIC |
| Age | 0.091 | 8353 | 8370 | 0.232 | 30871388 | 30871666 |
| + sex | 0.093 | 8340 | 8365 | 0.234 | 30789963 | 30790381 |
| + ethnicity | 0.094 | 8336 | 8394 | 0.241 | 30524687 | 30525662 |
| + IMD decile | 0.095 | 8326 | 8393 | 0.248 | 30241212 | 30242327 |
| + vaccine status | 0.145 | 7880 | 7989 | 0.297 | 28292202 | 28294012 |
| + LTC count | 0.224 | 7160 | 7277 | 0.364 | 25565855 | 25567805 |
| + polypharmacy | 0.225 | 7146 | 7272 | 0.369 | 25358599 | 25360689 |

Figure 1 visualises results from the Blinder-Oaxaca Decomposition analysis to decompose the mean difference in risk of severe COVID-19 between the autistic and general population groups into two components: the endowment effect, showing how much of the mean difference between the autistic and general population group is due to different distributions of the variable, and the coefficient effect, showing how much of the mean difference between the two groups is due to different influences of the same variable. As Figure 1 shows, the difference in non-age-adjusted probability of severe COVID-19 (0.066 in the general population, 0.033 in the autism group) can largely be attributed to the younger age profile of adults with autism.

**Figure 1.**
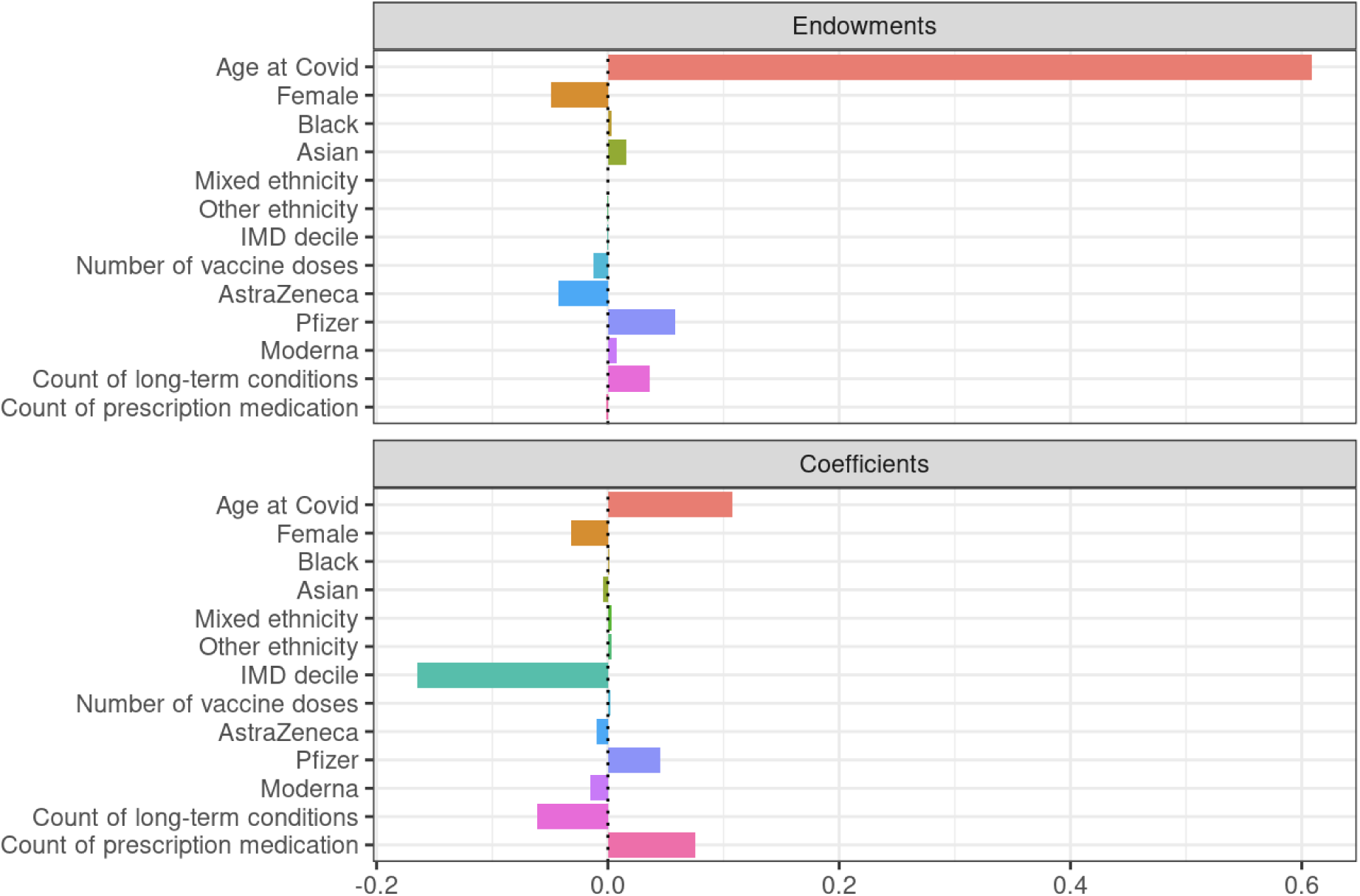
Visualisation of Blinder-Oaxaca Decomposition analysis.

## Discussion

This study aimed to explore COVID-19 in autism by investigating hospitalisation and mortality rates and exploring the association between demographic and clinical risk factors and their contributions to severe COVID-19 in autistic people. Previous studies investigating COVID-19 in autistic people produced conflicting results. Similar to a previous population-based study [34], our findings provide further evidence for the increased risk of hospitalisation (SIR 2020: 1.6, SIR 2021: 1.3) and mortality (SMR 2020: 1.52, SMR 2021: 1.34) of autistic adults due to COVID-19.

Exploration of demographic and clinical factors through regression modelling showed similar risk factors for the autistic and general population, with a few exceptions. Whereas all minority ethnicities were predictive of severe COVID-19 for the general population, only Asian/Asian British ethnicity was a significant predictor for the autistic population. This may be due to the different distribution of ethnicity between the two groups, with non-white ethnicities being less prevalent in the autistic population affecting sample size for those ethnicities (see Table 1). Similarly, female sex had a more protective effect for the general population than for the autistic population which may be explained due to the male bias in autism prevalence (43).

Number of COVID-19 vaccination doses only differed when not age-adjusted, however, the brands of vaccination had differing effects in the two populations. For both the general and autistic population, most people had received either Pfizer or AstraZeneca. Modelling of the predictors showed that while Pfizer was a significantly protective factor for severe COVID-19 for both populations, AstraZeneca was only significantly protective for the general population. Moderna was received the least by both populations and was found to be the most protective factor out of all predictors. There was no difference in the total number of vaccination doses received by either population.

The number of long-term conditions was the most important predictor of severe COVID-19 for the autistic population but not for the general population. Though the current study did not investigate individual associations between the long-term health conditions and COVID-19 hospitalisation and mortality, it is possible that this finding is due to the different clinical profile of the autistic population. Complex multimorbidity in general but particularly mental health conditions such as depression, dementia, anxiety and schizophrenia were more prevalent in the autistic study population. Previous research identified associations between increased risk of COVID-19 mortality and psychotic disorders, mood disorders and substance use disorders (9,10). The finding that the number of long-term conditions emerged as a significant predictor of severe COVID-19 in the autistic population, but not in the general population, underscores the intricate nature of multimorbidity in this group. However, this paper did not delve into the individual associations between long-term health conditions and COVID-19 outcomes. It is plausible that medication regimen, coupled with the presence of multimorbidity, may potentially lead to complex interactions that influence COVID-19 outcomes. Therefore, future research endeavours should aim to untangle the effects of medication regimen and the composition of multimorbidity in the autistic population on COVID-19 outcomes, warranting a separate and focused investigation.

Further, survival modelling showed that similarity in the number of prescriptions and the number of long-term conditions decreased the risk of severe COVID-19. In other words, low values of one variable paired with high values of the other were associated with higher risk of severe COVID-19. This may suggest that well-treated conditions and appropriate use of prescription medications may be a protective factor for severe COVID-19. Although this was found for both the autistic and general population, there is evidence to suggest that the health needs of autistic people are sometimes overlooked (44,45) or exacerbated by barriers in accessing medical care (46). Furthermore, despite similar effect sizes in the two populations, mortality rates were higher for the autistic population. Our findings highlight the importance and potentially far-reaching impact of addressing health inequalities experienced by autistic people.

Whilst polypharmacy was not included amongst the most important predictors in the hierarchical model for either the autistic or general population, further exploration suggests that this might be due to different effects of the medication class as we found, for both the autistic and general population, antipsychotic medication to be predictive of severe COVID-19 whereas antidepressant medication was protective. Furthermore, antipsychotic prescription was more predictive of severe COVID-19 in the general population whereas antidepressant prescription was more protective for the autistic population. Potentially, this could be due to differences in how these prescriptions are targeted in the two populations. For example, these medication classes are prescribed to autistic people on a more widespread basis with antipsychotics being prescribed in absence of comorbid psychiatric conditions (41) The finding of protective effects of antidepressant prescription corresponds to prior research suggesting lower infection rates and decreased COVID-19 severity in patients using antidepressants in the general population (47,48). Whilst there are contradictory findings, pooled effects seem to indicate a protective effect of antidepressants against severe COVID-19 (47).

Due to abovementioned targeting of prescription medications in the autistic population and potential of prescription medications to be used for a multitude of conditions, our study did not match prescribed medications to existing conditions as this would not be an accurate representation of the use of medications in the autistic population. As such, we are unable to ascertain whether there is a potentially mediating effect between medication and condition on severe COVID-19. For example, several studies provide evidence that psychiatric conditions such as schizophrenia, depression and anxiety have higher COVID-19 mortality rates (9,10). Results from the hierarchical modelling suggest that the long-term condition is indeed a more significant risk factor than polypharmacy. However, the classes of medication explored in this study can be prescribed for a multitude of conditions and, thus, were not linked to specific long-term conditions. In addition, there are concerns that there may be adverse reactions of prescription medication and COVID-19 treatments (20). Due to the unavailability of COVID-19 treatment data, this study was unable to ascertain whether prescription medication was taken during the hospital stay and whether there could be potential reactions with COVID-19 treatments provided. Additionally, exploring the effects of differing combinations of prescription medication(s) and long-term conditions was beyond the scope of the current study. However, our results suggest that this may be a viable target for future research.

### Strengths and limitations

This study has several strengths. The study makes use of a whole-country population of autistic individuals and the general population. This large sample size ensures a representative sample, thereby reducing sampling bias. The combination of the large sample size and the use of validated datasets further strengthens the validity and robustness of the results. Moreover, inclusion criteria for autism are based on clinical diagnoses, using both older and newer diagnosis codes relevant to autism. This allows us to better capture people on the spectrum and, thus, better represent the autistic population in England.

A limitation is that, in earlier stages of the pandemic, community incidence of COVID-19 is likely underreported as testing was not yet standard practice and availability of tests was limited. However, it is likely that testing rates do not differ between people with and without autism. Similarly, case fatality rates may be inflated due to lack of testing in these earlier stages. Additionally, whilst the dataset makes use of a whole-country population dataset, it should be noted that the final merged dataset contained a larger number of records than the recorded population size of England. This may be due to duplication of records, incorrect matching of NHS and death records, or people not registered as living in the UK receiving NHS treatment. However, effects on results reported in this study are likely to be minimal due to its use of rates and ratios, though caution should be exercised when interpreting absolute values.

Due to the use of autism identification through SNOMED CT diagnosis codes through primary care data, it is possible that the autistic study population does not fully capture the autistic population in England. Previous studies have indeed identified gaps in identification of autistic people over 30 years old within primary care (37). Moreover, it is possible that some autistic individuals are not registered with primary care and thus were not identified in this dataset.

## Conclusion

To conclude, our results show that autistic adults are at higher risk of hospitalisation or death due to COVID-19 as compared to the general population. Whilst demographic and clinical risk factors are similar for the autistic and general population, our results suggest that complex multimorbidity and prescription medication class may be of particular importance for the autistic population and should be further researched.

## Declarations

### Ethics approval and consent to participate

The North-East – Newcastle and North Tyneside 2 research ethics committee provided ethical approval for the CVD-COVID-UK/COVID-IMPACT research programme (REC No 20/NE/0161) to access, within secure trusted research environments, unconsented, whole-population, de-identified data from electronic health records collected as part of patients’ routine healthcare. The need for informed consent was waived by the North-East – Newcastle and North Tyneside 2 research ethics committee. All methods were carried out in accordance with relevant guidelines and regulations.

### Availability of data and materials

The data used in this study are available in NHS England’s Secure Data Environment (SDE) service for England, but as restrictions apply they are not publicly available (49).

The CVD-COVID-UK/COVID-IMPACT programme led by the BHF Data Science Centre (50) received approval to access data in NHS England’s SDE service for England from the Independent Group Advising on the Release of Data (IGARD) (51) via an application made in the Data Access Request Service (DARS) Online system (ref. DARS-NIC-381078-Y9C5K) (52).

The CVD-COVID-UK/COVID-IMPACT Approvals & Oversight Board (31) subsequently granted approval to this project to access the data within NHS England’s SDE service for England. The de-identified data used in this study were made available to accredited researchers only. Those wishing to gain access to the data should contact in the first instance.

## Acknowledgements

This work was carried out with the support of the BHF Data Science Centre led by HDR UK (BHF Grant no. SP/19/3/34678). This study makes use of de-identified data held in NHS England’s Secure Data Environment service for England and made available via the BHF Data Science Centre’s CVD-COVID-UK/COVID-IMPACT consortium. This work uses data provided by patients and collected by the NHS as part of their care and support. We would also like to acknowledge all data providers who make health relevant data available for research.

The authors would like to thank The Baily Thomas Charitable Fund for funding staff costs on this study.

## Data Availability Statement

For the purpose of open access, the authors have applied a Creative Commons Attribution (CC BY) licence to any Author Accepted Manuscript version arising from this submission.

## Footnotes

1 Throughout this paper we use identity-first language or neutral terms (‘autistic people’ or ‘people on the autism spectrum’) rather than person-first language as a result of the article by Kenny et al. (53) highlighting the preference for identity-first language by the majority of autistic people and their families.

