## Supplementary for "A Population-Based Cross-Sectional Investigation of COVID-19 Hospitalizations and Mortality Among Autistic People"

**Supplementary Material**

Percentage of the population to have a diagnosis of the 35 most common conditions in the UK. For the autistic population, both unadjusted and age-adjusted values are given.

| | Whole population | | | | | Positive COVID-19 test | | | | |
|---|---|---|---|---|---|---|---|---|---|---|
| | General population (n = 48,383,130) | Autistic population (n = 180,860) | | | | General population (n = 8,422,375) | Autistic population (n = 32,375) | | | |
| | | Adjusted | P-value | Unadjusted | P-value | | Adjusted | P-value | Unadjusted | P-value |
| Alcohol problems | 4.0 | 6.4 | <0.001 | 3.5 | <0.001 | 5.2 | 7.7 | <0.001 | 4.6 | <0.001 |
| Anorexia or bulimia | 0.2 | 0.5 | <0.001 | 0.5 | <0.001 | 0.3 | 1.0 | <0.001 | 0.9 | <0.001 |
| Anxiety & other neurotic | 4.9 | 12.6 | <0.001 | 9.3 | <0.001 | 7.5 | 17.3 | <0.001 | 12.3 | <0.001 |
| Asthma (currently treated) | 4.3 | 6.5 | <0.001 | 5.0 | <0.001 | 5.7 | 9.9 | <0.001 | 6.6 | <0.001 |
| Atrial fibrillation | 2.8 | 4.7 | <0.001 | 0.7 | <0.001 | 3.5 | 4.8 | <0.001 | 1.0 | <0.001 |
| Blindness and low vision | 0.8 | 1.5 | <0.001 | 0.4 | <0.001 | 1.1 | 1.5 | .02 | 0.7 | <0.001 |
| Bronchiectasis | 0.2 | 0.3 | .02 | 0.1 | <0.001 | 0.3 | 0.3 | .10 | 0.1 | <0.001 |
| Cancer - [New]Diagnosis in last five years | 8.9 | 8.6 | <0.001 | 3.5 | <0.001 | 12.2 | 10.3 | <0.001 | 4.4 | <0.001 |

| | | | | | | | | | | |
|---|---|---|---|---|---|---|---|---|---|---|
| Coronary heart disease | 3.6 | 6.5 | <0.001 | 1.1 | <0.001 | 4.9 | 7.6 | <0.001 | 1.7 | <0.001 |
| Chronic kidney disease | 0.1 | 0.2 | .002 | 0.1 | .001 | 0.1 | 0.2 | .11 | 0.1 | .23 |
| Chronic Liver Disease and Viral Hepatitis | 4.7 | 8.8 | <0.001 | 3.4 | <0.001 | 6.2 | 10.2 | <0.001 | 4.9 | <0.001 |
| COPD | 2.9 | 4.5 | <0.001 | 0.8 | <0.001 | 2.9 | 4.2 | <0.001 | 1.0 | <0.001 |
| Dementia | 3.9 | 10.0 | <0.001 | 4.8 | <0.001 | 5.6 | 13.6 | <0.001 | 6.8 | <0.001 |
| Depression | 4.6 | 14.0 | <0.001 | 12.5 | <0.001 | 6.6 | 18.6 | <0.001 | 14.6 | <0.001 |
| Diabetes | 9.8 | 15.4 | <0.001 | 4.1 | <0.001 | 11.0 | 16.6 | <0.001 | 5.0 | <0.001 |
| Diverticular disease of intestine | 3.0 | 6.3 | <0.001 | 2.1 | <0.001 | 4.3 | 8.1 | <0.001 | 3.1 | <0.001 |
| Epilepsy (currently treated) | 1.6 | 5.5 | <0.001 | 1.7 | .01 | 2.7 | 7.3 | <0.001 | 2.2 | <0.001 |
| Heart failure | 5.2 | 8.5 | <0.001 | 1.3 | <0.001 | 6.8 | 9.7 | <0.001 | 2.0 | <0.001 |
| Hearing loss | 0.9 | 2.1 | <0.001 | 0.4 | <0.001 | 1.2 | 2.4 | <0.001 | 0.6 | <0.001 |
| Hypertension | 8.0 | 11.6 | <0.001 | 2.3 | <0.001 | 9.1 | 12.5 | <0.001 | 3.0 | <0.001 |
| Inflammatory bowel disease | 1.4 | 2.8 | <0.001 | 1.1 | <0.001 | 2.1 | 3.9 | <0.001 | 1.7 | <0.001 |
| Irritable bowel syndrome | 0.9 | 1.9 | <0.001 | 0.8 | .001 | 1.2 | 2.6 | <0.001 | 1.2 | .45 |
| Migraine | 0.7 | 1.3 | <0.001 | 0.5 | <0.001 | 1.0 | 1.6 | <0.001 | 0.8 | <0.001 |
| Multiple sclerosis | 0.1 | 0.1 | .60 | 0.1 | .001 | 0.1 | 0.2 | .20 | 0.1 | .57 |
| Peptic Ulcer Disease | 1.4 | 2.8 | <0.001 | 0.9 | <0.001 | 1.9 | 3.6 | <0.001 | 1.4 | <0.001 |

| | | | | | | | | | | |
|---|---|---|---|---|---|---|---|---|---|---|
| Parkinson's disease | 0.3 | 1.1 | <0.001 | 0.1 | <0.001 | 0.4 | 1.2 | <0.001 | 0.2 | <0.001 |
| Prostate disorders | 0.8 | 2.3 | <0.001 | 0.4 | <0.001 | 1.1 | 2.8 | <0.001 | 0.6 | <0.001 |
| Psychoactive substance misuse (not alcohol) | 5.9 | 12.5 | <0.001 | 6.6 | <0.001 | 7.4 | 15.7 | <0.001 | 9.1 | <0.001 |
| Psoriasis or eczema | 0.6 | 1.4 | <0.001 | 0.5 | <0.001 | 1.0 | 2.1 | <0.001 | 0.7 | <0.001 |
| Peripheral vascular disease | 1.5 | 2.8 | <0.001 | 0.5 | <0.001 | 2.0 | 3.4 | <0.001 | 0.7 | <0.001 |
| Rheumatoid arthritis | 2.4 | 3.7 | <0.001 | 1.4 | <0.001 | 3.3 | 4.9 | <0.001 | 1.9 | <0.001 |
| Schizophrenia (and related non-organic psychosis) or bipolar disorder | 4.0 | 22.5 | <0.001 | 16.1 | <0.001 | 5.9 | 29.6 | <0.001 | 20.2 | <0.001 |
| Chronic sinusitis | 0.2 | 0.3 | <0.001 | 0.2 | .69 | 0.2 | 0.3 | <0.001 | 0.2 | .68 |
| Stroke & transient ischaemic attack | 4.2 | 9.8 | <0.001 | 4.3 | .32 | 6.1 | 13.4 | <0.001 | 6.3 | .08 |
| Thyroid disorders | 1.5 | 2.5 | <0.001 | 0.8 | <0.001 | 1.9 | 2.8 | <0.001 | 1.0 | <0.001 |
